# Compliance to food safety standards - Determining the barriers within the hotel industry

**DOI:** 10.1101/2023.12.13.23299917

**Authors:** Cynthia Esinam Segbedzi, Edward Wilson Ansah, Daniel Apaak

## Abstract

The safety of food served to customers is obligatory for all food service establishments for public health effects. However, workplace barriers have led to noncompliance with safety standards resulting in food contamination and outbreak of foodborne diseases. This study assessed the compliance of restaurant facilities to standards by the Food and Drugs Authority’s (FDA) code of standards, awareness and training on the code and the workplace barriers to compliance. The study involved 233 respondents, 205 food handlers, 10 managers, and 18 officers from the regulatory authorities, who were at work after the COVID-19 restrictions. Questionnaire was used to obtain data from the food handlers and face-to-face interview for managers/chefs and officers from the regulatory authorities. Data was analysed using frequencies and percentages, and thematic content analysis. Results revealed that majority of the hotels did not comply with the FDA’s code of standards on the provision of facilities. However, in segregating the items individually, 70% of the hotels had high compliance with the provision of proper storage facilities for raw and cooked food, 81.5% of the food handlers had in-service training, but this was not routine. Most of the food handlers were aware of the FDA’s code on hygiene for food service establishments, but only a few were trained on it. The food handlers reported unconducive working environment, poor monitoring and supervision, inadequate supply of equipment, time pressure and workplace policy as barriers to compliance with food safety standards. We recommend that hotel owners/managers should be adequately trained to provide the required training and supervision for food handlers, provide the needed tools/equipment to enhance work flow and safe food to consumers. Also, regulatory authorities are encouraged to conduct regular and effective monitoring/supervision to ensure adherence to standards, to improve the safety of food served to the public.

## Introduction

Access to safe and healthy food is a right to every human being for healthy living [1]. Safe and quality food therefore, contribute to food and nutrition security and sustainable development of a nation [2]. Yet, there is recurrent foodborne disease outbreaks worldwide due to noncompliance to safe food standards [3]. This is because food can be a latent avenue for infectious contaminants from preparation to consumption and it is more plausible in establishments where food is prepared on a large scale. Besides, the risk of getting infections is high, where asymptomatic food handlers prepare and serve foods to customers [4]. Foods contaminated by microbes are usually difficult to detect since these microbes are imperceptible and can cause illness [5,6]. The problem is exacerbated in countries where lack of food handling practices, inadequate food safety laws, weak regulatory systems, lack of financial resources to invest in safer equipment, and lack of food safety training are common leading to noncompliance to safe food standards [7,8,9]. Meanwhile, most foodborne diseases may be self-limiting yet, some can be very fatal leading to increased morbidity and mortality [5,10]. For instance, globally, over 600 million people die after consuming contaminated food [11]. The [1] estimates that over 47.8 million people fall ill with 128,000 hospitalizations and 30,000 deaths occur annually from foodborne diseases. However, the burden of foodborne diseases is estimated to be high in developing countries, including the Africa region [11].

In the Central and Western Regions of Ghana, foodborne disease infestation is on the rise and this may be due to asymptomatic food handlers that may be contaminating food and water [12]. The evidence is that, surveillance and epidemiological data indicate that poor food handling practices, inadequate infrastructural facilities and ineffective monitoring/supervision in food service establishments are linked to the causative chain of foodborne diseases [5,6,13]. Several previous studies [5,13,14,15,16,17] reported poor food safety knowledge, attitude and practices of food handlers however, few studies have been conducted on the barriers affecting compliance with food safety standards. Previous studies indicate that food preparation processes expose food to many contaminants and this needs to be addressed [18,19,20]. On the basis of this, it is imperative to investigate the barriers to food safety compliance by hotels in the Central and Western Regions of Ghana. Three research questions guided the study: (1) what is the level of compliance to FDA’s code of standards on the provision of restaurant facilities among hotels in the Central and Western Regions, Ghana? (2) What is the level of awareness and training on the FDA’s code of standard on food safety among food handlers at hotels in the Central and Western Regions, Ghana? and (3) What workplace barriers impede compliance to the FDA’s code of standard on food safety among food handlers at hotels in the Central and Western Regions, Ghana?

## Methods and Materials

A total of 21 hotels with restaurants located in the Central and Western Regions, which were in full operation after COVID-19 restrictions, were included in this study. This study surveyed 233 respondents consisting of 205 food handlers, 10 managers/chefs and 18 officers from the regulatory agencies (FDA, Environmental Health and sanitation Unit [EHS], & Ghana Tourism Authority [GTA]). The hotels included five 1star, nine 2star and seven 3star hotels. The census and purposive sampling techniques were used to select the participants and the hotels. The census procedure was used to include all hotels while purposive sampling was used to select the food handlers, managers/chefs and regulatory officers.

We secured ethical clearance (ID: UCCIRB/CES/2020/28) from the Institutional Review Board (IRB), University of Cape Coast. Besides, permission was obtained from the FDA, EHS, GTA, and the various hotel managers. The purpose of the research was clarified to the respondents and their anonymity and confidentiality were guaranteed. Informed consent forms were made available for the respondents who signed before taken part in the study [21].

The instruments were then distributed and collected two weeks after administration. Interviews for the food and beverage mangers/chefs and the regulatory officials were audio recorded and notes were also taken. The interviews were conducted at the offices of the regulatory officials and each session lasted about 40 minutes. Data collection commenced January 25 - May 20, 2021.

### Measures

The instruments included a questionnaire, an interview guide and an observation checklist. The instruments were adapted from related literature [6,16,22]. The questionnaire for food handlers was divided into four segments (A, B and C). Section ‘A’ surveyed the demographic characteristics of the respondents such as sex, age, level of education, training, and work experience. Section ‘B’ consisted of 16 items which responses ranged from “Yes” and “No”. This section dealt with the FDA code guidelines on compliance with food safety standards on the provision of facilities at hotels. Meanwhile, section ‘C’ consisted 11 items with a “Yes” and “No” responses measuring the perceived workplace barriers to compliance with food safety standards.

We used an observation checklist based on the FDA’s code for food service establishments [23], to conduct an on-site observation at all the 21 hotels to ascertain conformity of the facilities and kitchen environment to the code of practice. This covered areas like ventilation, lighting, wall/floors/ceilings, sanitary facility conditions, kitchen equipment and their locations, and storage facilities. Interviews were also conducted with the regulatory officials to explore their role in ensuring compliance with food safety standards at the hotels. Validity and reliability of the instrument were established through peer-review and pre-testing using four fast food restaurants in Winneba.

### Data analysis

To determine compliance to FDA’s code of standards, assessment of restaurant facilities was conducted by a member of the research team, observing the COVID-19 protocols (nose masking, hand-sanitizing, physical distancing, etcetera). The assessment was to examine the extent to which facilities (storage, work surfaces, water supply, sinks, washrooms, etc.) within the hotels complied with standards prescribed by FDA (FDA, 2013). Frequencies and percentages were used to analyse the observed data and compliance scores. The level of compliance was categorized into low <50, average 50-69 and high ≥ 70 [24]. The compliance scores were calculated using the equation below:

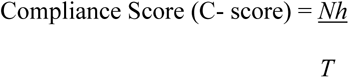

Nh = Number of hotels complying with a particular food safety standard.

T = Total number of hotels.

The self-reported barriers to compliance with food safety standards were analysed quantitatively via frequencies, simple percentages and tabulations.

## Results

The study involved 205 food handlers from 21 hotels, 10 managers/chefs and 18 officials from the regulatory agencies. Most of the food handlers were females (57.6%), while the rest (42.4%) were males. Many were between 21 and 30 years (68.8%), and 45.4% of the food handlers had 2 to 4 years working experience. Also, a greater proportion (81.5%) of the respondents have been trained on food safety.

### Compliance to food safety standards

The results indicated that half (50%) of the hotels did not comply with the FDA’s code on availability of facilities “Table 1”. However, in segregating the items individually, majority (70%) of the hotels had high compliance with the provision of proper storage facilities for raw and cooked foods. Furthermore, observation showed that a large percentage (80%) of the hotels have changing rooms and washrooms for food handlers. Moreover, a little over half (55%) of the hotel kitchens were well-ventilated, while some (55%) had poor work surfaces. Besides, a good proportion (65%) of the restaurants have their walls, ceilings and floors of light colour and made of easy to clean materials. Meanwhile, 75% of the hotels lacked probe thermometers for checking the doneness of cooked and held foods. Additionally, 75% of the doors to the food preparation/service areas and washrooms were not self-opening. Similarly, most (60%) of the hotels do not have a designated area for food handlers to eat, and 65% lacked separate sinks for handwashing in the kitchen. In addition, a large percentage (80%) of the restaurants were observed to have used same chopping boards for handling different foods, while a greater proportion (90%) did not have reminders or posters at vantage areas and above hand washing sinks to remind food handlers to wash their hands regularly, especially during the period of infectious disease outbreak like the Covid-19 pandemic.

**Table 1:** Compliance Scores and Corresponding Compliance Levels of Hotel Facilities to Food Safety Standards in Central and Western Regions.

| Compliance to food safety | Yes |  | No |  | Compliance Score | Compliance Level |
| --- | --- | --- | --- | --- | --- | --- |
|  | *F* | % | *F* | % |  |  |
| This hotel has adequate storage facilities for storing raw and cooked foods. | 14 | 70 | 6 | 30 | 70 | High |
| In this hotel thermometers are available for checking the temperature of cooked foods | 5 | 25 | 15 | 75 | 25 | Low |
| This hotel has spacious and well organized kitchen. | 7 | 35 | 13 | 65 | 35 | Low |
| This hotel has changing rooms for employees. | 16 | 80 | 4 | 20 | 80 |  |
| This hotel has a wash room that is conveniently located and readily accessible for food service employee. | 12 | 60 | 8 | 40 | 60 | Average |
| This hotel has a designated place for employees to eat their meals. | 8 | 40 | 12 | 60 | 40 | Low |
| This hotel has a designated sink for hand washing solely for food service workers with adequate handwashing materials. | 7 | 35 | 13 | 65 | 35 | Low |
| In this hotel appropriate materials are used for surfaces and they are easy to clean and maintain. | 12 | 60 | 8 | 40 | 60 | Average |
| Walls, ceilings and floors of this hotel is made of light coloured and easy to clean materials. | 13 | 65 | 7 | 35 | 65 | Average |
| The doors to washrooms, food preparation and service areas in this hotel is self-opening. | 3 | 15 | 15 | 75 | 15 | Low |
| In this hotel food handlers use separate chopping boards for handling foods. | 4 | 20 | 16 | 80 | 20 | Low |
| In this hotel, there is a sign/poster above every hand washing facility that reminds employees to wash their hands | 2 | 10 | 18 | 90 | 10 | Low |
| In this hotel there are waste bins with fitting lids. | 9 | 45 | 11 | 55 | 55 | Average |
| In this hotel the kitchen is well lit. | 12 | 60 | 8 | 40 | 60 | Average |
| This hotel has a well-ventilated kitchen. | 11 | 55 | 9 | 45 | 55 | Low |
| In this hotel, work surfaces are sound and in good condition (Rust free, smooth, not chipped). | 9 | 45 | 11 | 55 | 55 | Low |
| This hotel has adequate supply of water. | 8 | 40 | 10 | 60 | 60 | Low |

### Training of food handlers on food safety

The results showed that 81.5% of the food handlers reported they have ever been trained, and 58.5% received this training in 2019. However, in 2020, less than half (45.9%) of the food handlers received training “Fig 1”.

**Figure 1:**
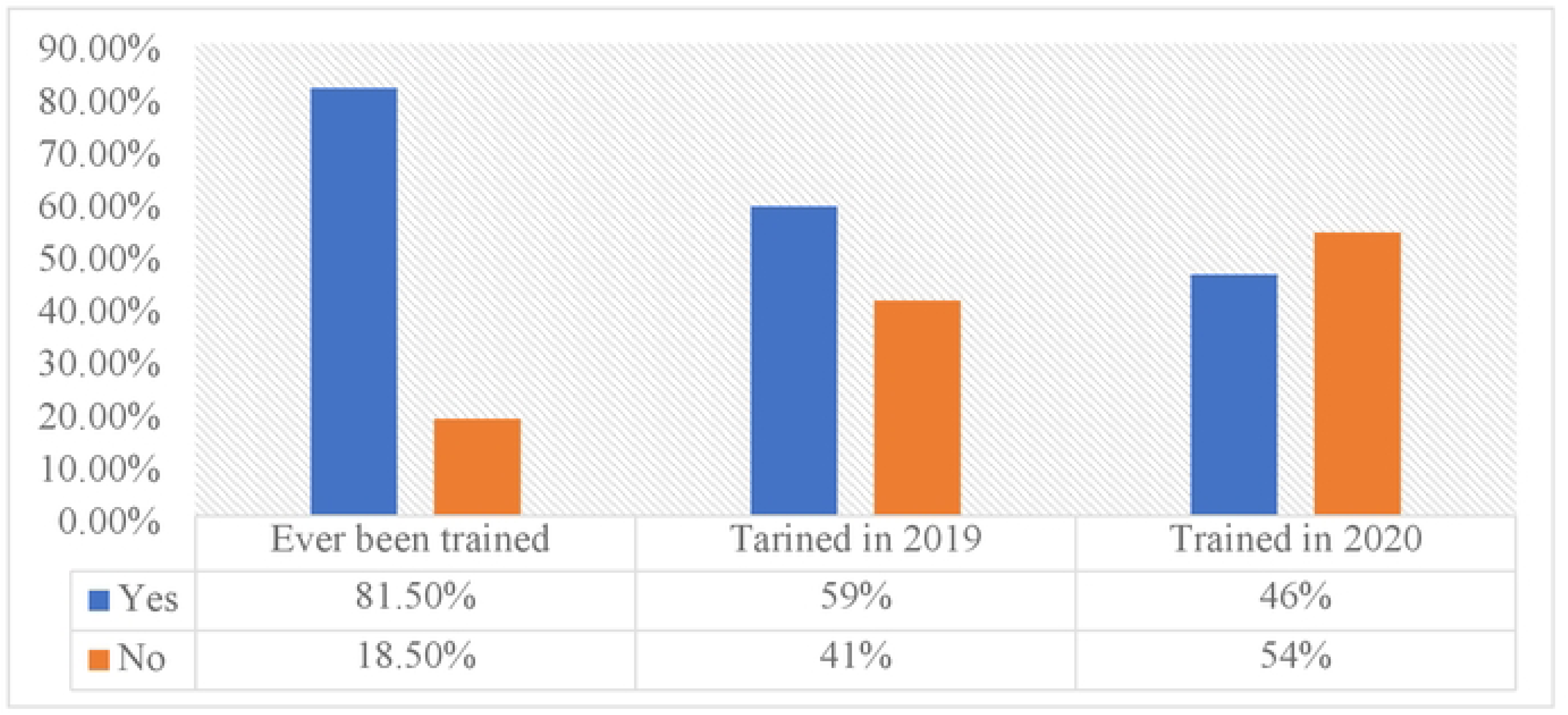
Food Safety Training among Food Handlers at Hotels in Central Western Regions.

### Training and awareness of the FDA code

The result further indicated that less than half (47%) of the food handlers were aware of the code, while only 40% reported to have been trained on these standards.

### Workplace barriers to food safety compliance

From Table 2, 66.3% of the food handlers indicated their workplace was not conducive for safe food preparation, posing as a barrier to safe food compliance. In contrast, more than half (63.4%) and 58.5% of the food handlers did not consider lack of motivation and inadequate knowledge on what they were expected to do as barriers to compliance. Meanwhile, 55.1% of the food handlers agreed that poor monitoring/enforcement are poor at their restaurants. In addition, 71.7% considered time pressure as barriers to safe food practices. Moreover, 55.6% and 63.9% of the food handlers indicated lack of equipment and their inconvenient locations are barriers to their food safety practices. Similarly, 63.9% and 78.5% of the food handlers agreed that food safety culture and employee welfare/workplace policies are barriers to safety practices. Besides, 64.9% of the food handlers indicated forgetfulness could compromise compliance with food safety standards in their hostels.

**Table 2:** Barriers to Food Safety Practices among Food Handlers at Hotels in Central and Western Regions.

| **Barriers to Food Safety compliance** | **Yes** |  | **No** |  |
| --- | --- | --- | --- | --- |
|  | *F* | % | *F* | % |
| Unconducive work environment in terms of structure (physical layout). | 136 | 66.3 | 69 | 33.7 |
| Lack of motivation. | 75 | 36.6 | 130 | 63.4 |
| Inadequate knowledge on what one is expected to do. | 85 | 41.5 | 120 | 58.5 |
| Lack of training and education on appropriate food safety practices. | 109 | 53.2 | 96 | 46.8 |
| Small working space | 112 | 54.6 | 93 | 45.4 |
| Poor monitoring and enforcement by supervisors and regulatory authorities. | 113 | 55.1 | 92 | 44.9 |
| Time pressure or busy work schedules. | 147 | 71.7 | 58 | 28.3 |
| Lack of necessary equipment and resources. | 114 | 55.6 | 91 | 44.4 |
| Inconvenient location of equipment such as sinks. | 131 | 63.9 | 74 | 36.1 |
| Food safety culture. | 131 | 63.9 | 74 | 36.1 |
| Employee welfare and workplace policies | 161 | 78.5 | 44 | 21.5 |
| Forgetfulness | 133 | 64.9 | 72 | 35.1 |

## Discussion

The aim of this research was to examine the barriers to food safety compliance by hotels in the Central and Western Regions of Ghana. The findings revealed that a greater number of the hotels have high compliance levels for the provision of only storage facilities, but they neglected the other facilities. Besides, majority of the food handlers had in-service training, but this was not a routine practice. Meanwhile, many of the food handlers reported they work in an unconducive work environment, with time pressure, lack of equipment/inconvenient location and small work space. These factors pose as workplace barriers to compliance with food safety standards in these hotels.

### Compliance with food safety standards

Our finding revealed that most (70%) of the hotels have facilities for storage (refrigerators, freezers, chillers and dry storage areas), changing rooms, washroom solely for food handlers to use, well-ventilated kitchens with the walls, floors and ceiling in good condition. This agrees with the results of [25] who also observed decreased violations in compliance regarding cold and dry storage facilities, personnel facilities, utensils and equipment in the Greater Accra Region of Ghana. Thus, the availability of these facilities helps food handlers to prevent contamination, spoilage of food and transmission of foodborne infections.

Unfortunately, most of the hotels lack thermometers for checking the temperature of cooked foods, a place for food handlers to eat their meals, handwashing sinks, separate chopping boards for different foods and no reminders on top of handwashing sinks. The lack of these facilities suggests low level of compliance to standards that could result in the exposure of consumers to foodborne diseases. Unfortunately, this does not align with the FDA’s code of standards on the provision of facilities at food service establishments. The consequence is that this is likely to hinder safe food handling practices and expose consumers to health risks [15,22,26]. Thus, the two regions may continue to record high foodborne infection rates.

### Training of food handlers on food safety

The finding further showed that most of the food handlers received in-service training on the FDA’s code of standard, though not regularly (see Figure 1). This may pose a barrier to compliance because there are inadequate opportunities for the food handlers to update their knowledge and skills needed for safe food handling. Then, it becomes important to increase employee’s awareness of workplace practices through consistent training or workshops to increase their motivation and confidence to improve adherence to proper food handling practices to minimize food contamination and its associated hazards on consumers’ health [14]. In addition, training should target all food handlers, be regular, tailored to specific function and level of knowledge, eliminate irrelevant information, use appropriate media to disseminate information and in the preferred language of the food handlers [18].

*For instance, manager (CWR2) responded this way:*

> *We organize training but we don’t have specific days for it. Our training is on needs basis. That is when there is customer complaint or we observe a bad practice among the food handlers. Sometimes we combine training and work due to time constraints. We would prefer to have the regulators add their bit from-time-to-time to reinforce what we do.*

*A female officer (TCR2FG2) also remarked:*

> *In fact, as regulators, one of our core functions is to inspect to prevent violations and train hotel staff. However, we are resource constrained. We expect the hotels to be a bit more committed to training their employees, but the hotel industry is one with a high employee turnover. So, they feel it’s a waste to commit resources into training staff only for them to leave in a few months.*

These findings are consistent with the observations from previous studies [16,18,27] indicating that most food handlers do not receive regular training resulting in increased food safety violations and risk to public health. We are of the view that the reduction in the number of food handlers trained in 2020 is attributable to the outbreak of the COVID-19 pandemic which resulted in restrictions/ban on hotel activities and staff lay-offs.

### Training and awareness of the FDA code

To ensure that food handlers complied with the FDA’s code on food safety standards, it is mandatory for food handlers to be aware of the regulatory requirements and be trained to manage any condition that has the potential for risk within the food service environment. We found however, that not many of the food handlers were aware of this code of practice and only a few of them had training on such codes (see Figure 1). Although, many of the food handlers have high level of education, the lack of awareness and training on their specific roles may result in noncompliance with food safety standards. Meanwhile, the food safety standards are numerous and require training to understand current trends and requirements within the industry [28,29]. Therefore, it is important that managers/owners or supervisors are equipped with adequate knowledge and resources to be in position to train their workers as stipulated in the FDA’s code of standards for food hygiene practices at food service establishments. A regulator (TCR1FG1) observed:

> *Most of the time we only stress on managers/owners train the food handlers. Some of the managers are highly educated but do not have the required knowledge in food safety. So, we think the managers also need training so as to impart the required knowledge and skills to the food handlers.*

Thus, the lack of awareness and training on the code of practice is likely to be a serious barrier in complying with food safety standards. This is because food handlers are likely to engage in poor food safety practices that would expose consumers to myriad of health conditions [9,14,17].

### Workplace barriers to food safety compliance

The finding showed that many of the food handlers perceived that their work settings areunconducive with high level of time pressure, lack of equipment/inconvenient location, small working space, unpleasant food safety culture and employee welfare which serve as major barriers to compliance with food safety standards at hotel restaurants. A male chef (CWR1l) llamented:

> *Most of the hotel kitchens are not properly designed with food safety in mind. As I speak with you now, we are planning to restructure our kitchen to make it more convenient and comfortable to work in.*

A manager (CRW3) also observed:

> *I think hotel owners need to do broader consultation with architects and chefs to provide the technical ideas before construction of their facilities. This will enable the facilities to have the fine details to meet the standard requirements of a hotel kitchen. Sometimes you visit a hotel kitchen and the size is just too small for even a household usage. This makes it difficult to install the needed equipment to facilitate efficient workflow and also prevent cross-contamination.*

*Another male chef (CWR2) bemoaned:*

> *We are constrained in terms of resources in this hotel. Most of our equipment are broken down and a process which could be undertaken within a few minutes takes hours to complete.*

These admissions have dire implications because they could lead to increased risk of food contamination and poisoning of consumers increasing the burden of food-related illnesses. Therefore, awareness of these barriers and their effects on public health will provide the motivation for managers/regulators and employees to improve food safety practices. For instance, the observations of [6,25] are that most restaurant kitchens are the last to be designed, and some are residential facilities which were converted to restaurants. Therefore, the layout of such facilities does not conform with standards and do not also have sufficient space to install equipment needed for easy flow of work, efficiency and safety of food.

Furthermore, although all the barriers identified could hinder compliance to safe food standards, lack of training, lack of equipment, poor monitoring and supervision and forgetfulness are of greater concern. These could result in increased level of work-related stress leading to poor safe food practices which inadvertently endanger public health. This calls for a strong regulator activity, which seems relegated at these hotels.

A female regulator (TCR2FG2) decried:

> *We wish to go for visits and inspections at least thrice in a year, but we lack the personnel and the resources to carry it out. The number of personnel in-charge for ensuring enforcement and compliance is inadequate. Besides, some of the hotels are located far away from town. Sometimes we have to go on visits in hired taxis at our own expense.*

A male participant (TCR3FG3) bemoaned:

> *Sometimes we try to put human face to some of the activities of the various facilities, so, it may seem as if we are not doing our work. If we want to strictly enforce the regulations most facilities will be closed down. At times we go on visits unannounced, but usually we inform the facilities of our visit to afford them time to put their house in order. Yet, we still observe a number of food safety violations.*

*Another female participant (TCR4FG2) responded:*

> *You see……our work as regulators would have been much easier but there is no collaboration among the agencies. We do not share information, meanwhile we are regulating the same facilities.*

The finding of the current study bears similarity with several previous studies [6,13,15,26,30] instance, a study carried out by [31] to examine compliance to safe food standards among fast-food business in Jamaica revealed that food safety regulators could be considered as barriers to compliance due to inconsistencies in their actions to ensure adherence to national food safety codes.

Many of the food handlers may be working against time pressure, which could make them forget to follow laid down procedures. To minimize this, employees and managers can examine their work processes and alter or restructure work processes to increase productivity. For example, strategies such as location of work stations, ergonomic design of equipment, or training focused on smart working, will help increase compliance to food safety standards during meal preparation and service to protect consumers’ health [6,15,30,26]. Such restaurant may be said to have a good food safety culture that influences compliance with food safety standards. Food safety culture demonstrates how owners, managers and employees think and act on daily basis as well as their understanding, adopting and committing to measures to ensure food served to consumers is safe [15]. According to [32], employees specified that it is crucial for food service managers to be keenly involved within the operations of the organization, by demonstrating exemplary leadership, create supportive environment and regularly stress on the importance of food safety even during busy hours, to ensure that proper food safety behaviour is being practiced.

A good number of the food handlers indicated employee welfare and workplace policies pose as barriers to their food safety compliance. Workplace policies serve as the basis for building organizational culture. If workplace policies are unfavourable or not employee-friendly, it may affect work performance and compliance to safety standards. This implies that any policy which does not factor employees needs like their career progression, physical and mental health, may impact productivity negatively and lower safe food practices. This has implications for public health because of effect on increased contamination of food resulting in outbreak of foodborne diseases such as cholera, typhoid, dysentery, among others. For instance, previous studies [6, 33, 34] involving chefs, highlighted the practice of working for long hours in the kitchen as having an adverse effect on the physical and mental health of the chefs. It was recommended that management review their policies and promoted good work and motivating environment to increase attention to details, prevent high turnover of key employees, absenteeism and strikes or sabotage.

## Limitations

This study had some limitations. The COVID-19 restrictions placed limitation on the number of hotels included in the study. It would have been prudent to interview the food handlers in addition to the managers and regulators to properly triangulate the issues. However, we are confident that the information gathered with the study is relevant and can be generalised for the large population.

## Conclusions

We conclude that many of the hotels did not comply with the FDA’s code of standard on the provision of restaurant facilities to ensure safe food handling practices. Also, a large percentage of the food handlers received in-service training, but this was not routine. Several workplace barriers such as unconducive work environment, lack of training, time pressure, poor monitoring/enforcement, lack of equipment, poor food safety culture, forgetfulness, and workplace policies were reported by the food handlers to influence their low level of compliance to food safety standards. The cases of cholera and diarrheal diseases are likely to continue and increase in the Central and Western Regions of Ghana if compliance to food safety protocols are not enforced to the later. Thus, owners/managers of hotels owe it a responsibility to procure the required tools/equipment to ensure smooth work flow, proper training of food handlers and effective supervision to curb the risk of food contamination and safeguard public health. Moreover, the regulatory authorities need to increase the number of visits to the various hotels for proper monitoring and supervision.

## Ethical Approval

Ethical approval for the study was obtained from the Institutional Review Board (IRB) at University of Cape Coast, Ghana (UCCIRB//CES/2020/28) and medical screening and certification of first author by Environmental Health and Sanitation Unit of Cape Coast, Ghana (Cert. No: 1,057).

## Data Availability

All data pertaining to this research are attached to the manuscript

## Acknowledgement

We are grateful Mr. Botha Nkosi Nkosi of Air Force Medical Services, Takoradi, for proofreading the draft manuscript.

